# Quantification of amyloid protein enrichment by mass spectrometry improves amyloidosis typing

**DOI:** 10.1101/2023.04.07.23285834

**Authors:** Sylvaine Di Tommaso, Bertrand Chauveau, Cyril Dourthe, Jean-William Dupuy, Frédéric Saltel, Brigitte Le Bail, Anne-Aurélie Raymond

**Author notes:** These authors have contributed equally (Sylvaine Di Tommaso, Bertrand Chauveau, Brigitte Le Bail, Anne-Aurélie Raymond).

## Abstract

**Introduction:** Amyloidosis typing is crucial to determine the best therapeutic strategy for patients. Since conventional histological techniques often fail, the identification of amyloid precursors by mass spectrometry became the new standard. However, without quantification, selecting the amyloid precursor from proteins that may be ubiquitous under non-pathological conditions may be equivocal. Therefore, we quantified protein enrichment in amyloid deposits to improve typing.

**Methods:** Protein enrichment was measured by extracted ion chromatogram based label free quantification by comparing a microdissected amyloid area with a non-amyloid area. We assessed the discrimination ability of candidate precursors with this approach compared to the two practiced identification methods.

**Results:** As proof of concept, we selected seven cases, 5 typical of the most common amyloidosis subtypes and typed by immunostainings, 2 inconclusive after immunohistochemistry. Proteins associated with amyloid deposits were identified in all samples confirming the pathology. When the routine clinical mass spectrometric identification techniques allowed unambiguous conclusions for 2/3 of 7 cases, quantification of the enrichment ratio in the amyloid deposit allowed unambiguous precursor selection in all cases.

**Conclusion:** Quantification of precursor enrichment in amyloid deposits is a promising optimization for amyloidosis typing. Incorporated into routine clinical processes, it will improve patient care in difficult diagnostic situations.

## Introduction

The amyloidoses are a rare group of heterogeneous diseases resulting from the deposition of amyloid, *i.e*. a fibril-organized deposit based on twisted stacks of protein layers in ß-sheet structure [1,2], generally in extracellular space. Depending on the location of the deposits and the cause of the protein misfolding, amyloidosis can be either systemic or localized, acquired or hereditary. Amyloid deposits are composed of a specific protein precursor admixed with common additional components such as heparan sulfate proteoglycan and serum amyloid P component [2]. Amyloid precursors are listed by the International Amyloidosis Society constituting the basis of the nomenclature of amyloidosis. Their number is constantly increasing. In 2018, 36 precursors were recognized [3] compared to 42 today [4]. Common protein precursors of systemic amyloidosis include immunoglobulin light chain kappa and lambda (AL amyloidosis), transthyretin (ATTR, either acquired or hereditary) and serum amyloid A (AA), all four representing about 90% of amyloid cases in routine diagnosis [5]. Treatment of amyloidosis depends on the protein precursor, the localized or systemic nature of the deposits and/or the associated disease. It can range from chemotherapy, immune therapy or autologous stem cell transplant for AL amyloidosis, to suppression of inflammation for AA amyloidosis and siRNA therapy or even to liver transplantation in some cases of hereditary ATTR amyloidosis [6]. Amyloidosis typing is as such of utmost importance for patient medical care and for genetic counseling.

The diagnosis of amyloidosis can be suspected by clinicians or made incidentally by pathologists. The definite diagnosis requires tissue examination of an affected tissue or organ by a pathologist. Accessory salivary glands, subcutaneous abdominal fat and colonic mucosae are often involved in most common amyloidosis and represent easy targets for the biopsies. Amyloid deposits have a characteristic histological appearance, due to its affinity for Congo red stain and its yellow to green birefringence under polarized light. The typing of amyloidosis can be made in routine practice by immunofluorescence from frozen samples when fresh tissues can be obtained. Furthermore, except for reference centers, few antibodies are available. Chromogenic immunohistochemistry from formalin-fixed and paraffin-embedded (FFPE) samples is easier to perform, but it is prone to background noise, especially for light chains and transthyretin. Moreover, like immunofluorescence, specific antibodies targeting rare subtypes of amyloidosis are often not available, and samples are often too small to test many proteins. Immunoelectron microscopy is an alternative, but shares the same limitations in antibodies availability, are limited to few expert centers and requires a dedicated sample, rarely available in practice.

Hence, over the last decade, mass spectrometry-based proteomics from FFPE samples has emerged as the new gold standard for amyloidosis typing, and remains for the moment the pioneer clinical application of proteomics by mass spectrometry for surgical pathology laboratories [5,7]. It is especially indicated when typing by routine techniques is equivocal, conflicting with clinical data or non-informative [8]. The Mayo Clinic and the UK National Amyloidosis Centre are currently the two major centers of amyloidosis typing by mass spectrometry, having reported hundreds to thousands of cases with high performances in amyloidosis typing [5,7,9]. For example, Rezk *et al*. reported 80% of informative amyloidosis typing when the IHC technique was not [10] and a 100% rate of identification of an amyloid precursor has been reported by the Mayo Clinic [5,9]. Both centers share a common bottom-up proteomics workflow: laser microdissection of amyloid deposits stained by Congo red, extraction and digestion of the proteins by trypsin before peptide analysis by liquid chromatography and tandem mass spectrometry (LC-MS/MS). They both confirm the amyloid nature of the sample by the identification of amyloid-associated proteins, which includes serum amyloid P component, apolipoprotein E and apolipoprotein A-IV [5,7]. However, for precursor identification, the Mayo Clinic selects the precursor with the highest number of MS/MS sequencing runs (peptide spectrum matches, PSM) for specific peptides of a given protein, which indirectly correlates with the amount of protein in the sample. On the other hand, the UK National Amyloidosis Center selects the precursor protein with the best identification score obtained with the Mascot algorithm [7], which is classically used to evaluate the statistical confidence level of protein identification [11,12]. However, problems remain in identifying precursors, as proteomics frequently detects multiple potential precursors in the same sample, sometimes with PSM or Mascot scores at similar levels. Noteworthy examples include samples with few amyloid deposits, with blood contamination or with a protein precursor that is naturally present in tissue such as apolipoprotein A-I, A-IV, immunoglobulins and transthyretin [7,10,13]. As another example, when implementing PSM-based mass spectrometry typing of amyloidosis, an Australian team reported up to 40% AL amyloidosis with other identified secondary precursors, sometimes with PSM of similar magnitude [14].

The identification of a protein by mass spectrometry depends on its intrinsic physicochemical properties and thus on the ionization capacities of its tryptic peptides. Thus, a protein with good ionization capacity and in low quantity can be better identified than an abundant protein with poor ionization properties. Furthermore, identification by MS is also highly dependent on the number of tryptic peptides generated and thus on the sequence length of each protein. Therefore, relative protein quantification by mass spectrometry only compares the relative intensities of peptides of the same sequences with the same modifications and charge state. Although it is logical to think that a more abundant protein will be better identified and will benefit from more MS/MS sequencing, by a conventional analytical approach it is not accepted to compare the identification scores, number of MS/MS spectra, nor relative abundances between different protein species. Therefore, comparing the relative intensities of different protein species is not considered by conventional proteomic approaches as quantification [15].

Targeted quantitative mass spectrometry approaches had been proposed to improve the typing of amyloidosis. They consisted in measuring the amount of amyloid precursors in the deposits by adding standard isotopically labeled peptides. Ogawa *et al*. carried out a proof of concept [16] and Park *et al*. showed that targeted quantification displayed high sensitivity and specificity for amyloidosis typing compared to the standard mass spectrometry identification, even though it was only developed for the 3 main common types of systemic amyloidosis (AL, AA and ATTR) [17]. These pseudo-absolute quantity measurement methods, which require advanced expertise in mass spectrometry and specific acquisition methods, have not yet been integrated into routine clinical analyses.

Label free quantification is classically used in exploratory basic research projects and emerging clinical applications [18, 19]. Label free quantification is based on extracting the relative abundance of specific precursor peptides from the ion chromatogram (XIC) using chromatographic retention time and m/z values as specific coordinates for each ion. It has the advantage of being able to extract the relative abundance even if it is not systematically identified provided that the peptide has been sequenced by MS/MS at least once in at least one sample. Label free quantification enables a comparison of protein relative abundances between one sample and another, making it particularly suitable for the quantification of protein enrichment in amyloid deposits. There is to date no study describing the quantification of amyloid precursor enrichment ratios by label free approach for amyloidosis typing from FFPE samples. With this study, we would like to propose to implement in routine practice the enrichment quantification to make amyloidosis typing more robust.

## Materials and methods

### Study population

This study relied on a retrospective inclusion of selected patients with an amyloidosis diagnosis performed at the Bordeaux University Hospital from 2013 to 2022, with consistent clinico-biological data. Archived formalin-fixed and paraffin embedded (FFPE) samples with sufficient remaining material were included. Main clinical, biological and pathological characteristics were retrieved from the institutional medical software. Congo red staining was performed for amyloidosis diagnosis in all samples and amyloidosis typing was performed by routine immunohistochemistry targeting kappa and lambda light chains (clone A21-Y and K22-Y from Clinisciences), serum amyloid A (clone MC 1 from Dako), transthyretin (polyclonal from Dako) and β2-microglobulin (polyclonal from Genetex). All immunostains were performed using an Omnis automate from Dako Agilent (EnVision Flex/horseradish peroxidase for signal amplification). All reagents were provided by Dako/Agilent. No frozen tissue was available for any case. For each case, the abundance of amyloid deposits was semi-quantitatively assessed by a pathologist in a 1 to 5 scale, where 5 represents a sample nearly exclusively affected by amyloid deposits.

The study was conducted according to the guidelines of the Declaration of Helsinki and was approved by a local ethic committee for the protection of persons (reference number CER-BDX-2023-02).

### Microdissection of selected area and sample preparation

For each case, Hematoxylin eosin saffron and Congo red stains were used to determine the areas of interest to be dissected. Non-amyloid (NA) and Amyloid (A) areas were selected under the supervision of a pathologist (either B.LB. or B.C.). Between 0.1 and 1 mm^2^ of tissue was microdissected from a FFPE 5 μm-thick section with a PALM type 4 (Zeiss) laser microdissector.

Laser microdissected sections were collected and immerged in a 50 mM Ammonium bicarbonate buffer. Fragments were heated at 90°C for 120 min with occasional vortexing. Samples were reduced in 10mM dithiothreitol and alkyled in 100mM iodoacetamide before digestion into tryptic peptides overnight and analyzed by liquid chromatography and tandem mass spectrometry (LC-MS/MS).

### Mass spectrometry analysis

NanoLC-MS/MS analysis was performed using an Ultimate 3000 RSLC Nano-UPHLC system (Thermo Scientific, USA) coupled to a nanospray Orbitrap Fusion™ Lumos™ Tribrid™ Mass Spectrometer (Thermo Fisher Scientific, California, USA). Each peptide extracts were loaded on a 300 μm ID x 5 mm PepMap C18 precolumn (Thermo Scientific, USA) at a flow rate of 10 μL/min. After a 3 min desalting step, peptides were separated on a 50 cm EasySpray column (75 μm ID, 2 μm C18 beads, 100 Å pore size, ES903, Thermo Fisher Scientific) with a 4-40% linear gradient of solvent B (0.1% formic acid in 80% ACN) in 57 min. The separation flow rate was set at 300 nL/min. The mass spectrometer operated in positive ion mode at a 2.0 kV needle voltage. Data was acquired using Xcalibur 4.4 software in a data-dependent mode. MS scans (m/z 375-1500) were recorded at a resolution of R = 120000 (@ m/z 200), a standard AGC target and an injection time in automatic mode, followed by a top speed duty cycle of up to 3 seconds for MS/MS acquisition. Precursor ions (2 to 7 charge states) were isolated in the quadrupole with a mass window of 1.6 Th and fragmented with HCD@28% normalized collision energy. MS/MS data was acquired in the Orbitrap cell with a resolution of R=30000 (@m/z 200), an standard AGC target and a maximum injection time in automatic mode. For protein identification, Mascot 2.5 algorithm through Proteome Discoverer 2.5 Software (Thermo Fisher Scientific Inc.) was used in batch mode by searching against the UniProt Homo sapiens database (79 071 entries, Reference Proteome Set, release date: January, 2022) from http://www.uniprot.org/website. Two missed enzyme cleavages were allowed for the trypsin. Mass tolerances in MS and MS/MS were set to 10 ppm and 0.02 Da. Oxidation of methionine and acetylation of lysine were searched as dynamic modifications. Carbamidomethylation on cysteine was searched as static modification. Raw LC-MS/MS data were imported in Proline Studio for feature detection, alignment, and quantification [20]. Proteins identification was accepted only with at least 2 specific peptides with a pretty rank=1 and with a protein False Discovery Rate (FDR) value less than 1.0% calculated using the “decoy” option in Mascot. Label free quantification of MS1 spectra by extracted ion chromatograms (XIC) was carried out with parameters indicated previously [21]. The mass spectrometry proteomics data have been deposited to the ProteomeXchange Consortium via the PRIDE [22] partner repository with the dataset identifier PXD039814.

### Bioinformatical and statistical analysis

Statistical analyses and plots were performed using R, version 4.2.2 [23]. Plots were performed using the ggplot2 package and the ggpubr package. Spearman’s rank correlations were performed using the stats package. To assess the confidence in data interpretation, we defined a confidence score for each approach, expressed as a ratio (score of the first precursor divided by the mean of the next 2 scores). As all three methods of mass spectrometry-based amyloidosis typing hypothesized that, from all identified and/or quantified proteins, a protein precursor with a top score will emerge (PSM, Mascot or abundance ratio), confidence in results interpretation can be summarized, for each approach, as the relative difference between the protein precursor with the best score compared to the next two scores.

## Results

### Clinical and biological characteristics of the patients

To define our test panel of 7 cases, we selected cases without diagnostic ambiguity, focusing on the most common amyloidosis types. For 5 of them, immunohistochemistry (IHC) was informative, and for two of them IHC was not informative but the associated clinical data of the patient strongly suggested the amyloid precursor determination. Of these 7 patients, 6 were cases of systemic amyloidosis (case numbers 1 to 6) and 1 was a case of localized amyloidosis (case 7). Mean age of included patients was 67 years old (range 47-83). Available samples consisted of a biopsy for 3 cases and a surgical specimen for 4 cases, from various organ or tissue origin (**Table 1**). Amyloidosis typing based on clinicopathological findings were as follows: 1 AL lambda, 1 AL Kappa, 1 Aβ2M, 1 AA and 1 ATTR. In two cases (cases 4 and 7), IHC was non-informative. Case 4 had an IgM Kappa monoclonal gammopathy with an incidental finding of amyloid deposits in a duodenal ampulloma resection. The other case (case 7) had a previous biopsy that favored kappa light chain deposits by IHC. Main clinicopathological characteristics are summarized in **Table 1**.

**Table 1:** Relevant clinical data of the included patients

| Case No. | Age Range (years) | Gender | Sample (surgical specimen, biopsy) | Site | Amyloid subtype by IHC | Associated condition |
| --- | --- | --- | --- | --- | --- | --- |
| 1 | 65-70 | F | Surgical specimen (gangrene) | Perineum | β2-microglobulin | End-stage renal disease with dialysis for 40 years |
| 2 | 75-80 | M | Biopsy | Minor salivary gland | Light chain Lambda | Multiple myeloma (IgG Lambda) |
| 3 | 45-50 | F | Biopsy | Rectum | Serum amyloid A-1 protein | Familial Mediterranean fever. Hemodialysis |
| 4 | 80-85 | M | Surgical specimen | Duodenum (ampulloma) | Non-informative | IgM monoclonal gammopathy (kappa). Suspicion of low-grade digestive lymphoma. |
| 5 | 55-60 | M | Surgical specimen | Liver (recipient) | Transthyretin | Transthyretin-related hereditary amyloidosis |
| 6 | 65-70 | F | Biopsy | Bone marrow | Light chain Lambda | Monoclonal gammopathy (IgG Lambda) |
| 7 | 65-70 | F | Biopsy | Larynx | Undefined from this sample. A prior biopsy favored Kappa light chain deposits. | Localized amyloidosis |

### Quantification of protein enrichment in amyloid deposits and comparison with routine identification selection methods

For each case, two regions of interest of the same surface (1 mm^2^) were isolated by laser microdissection (**Figure 1**), one with amyloid deposits (**Figure 1b**) and the other without morphological amyloid deposits (**Figure 1c**). These regions were previously annotated on a Congo red staining by an experienced pathologist (**Figure 1d and e**). After microdissection (**Figure 1f**), proteins were extracted and digested, and proteolytic peptides were analyzed by high resolution tandem mass spectrometry (LC-MS/MS) (**Figure 2a**). In the amyloid area, identification of at least two of the three proteins associated with amyloid deposition (serum amyloid P-component/APCS, apolipoprotein E/APOE, and apolipoprotein A-IV/APOA4) was used as an initial quality control. For amyloidosis typing, the two methods routinely used in the expert centers are (i) the PSM, the protein precursor with the most peptide spectra matches (PSM, number of MS/MS spectra) in the amyloid area (A) and (ii) the Mascot score, the precursor with the highest Mascot identification score in the amyloid zone (A). We compared these methods to (iii) the quantification of the enrichment ratio by a label free approach: the precursor with the highest enrichment ratio calculated by comparing its relative abundance between the amyloid area (A) and the non-amyloid area (NA) (**Figure 2b**). A selection is first made from the 42 known amyloid precursors, which means that among the identified proteins we do not consider proteins that have never been associated with amyloidosis in first intention. Overall, in all analyzed cases, we identified between 4 and 34 amyloid precursor proteins per case. The quantification of the protein enrichment in the deposit (ratio A/NA) revealed that several precursors could be enriched and allowed us to isolate the precursor with the highest enrichment rate. Some ubiquitous amyloid precursors were even more present in the non-amyloid tissue than in the deposit (ratio A/NA ≤ 0.5). These results demonstrate the interest of providing quantity information beyond the simple identification of precursors (**Figure 2b**). We then focused on each of the analyzed cases. As a first example, case 3 is an inflammatory AA amyloidosis that was previously well characterized by IHC (**Figure 3a**). Amyloidosis-associated proteins (ApoE, ApoA4 and APCS) were identified and were enriched in the deposit (**Figure 3b**). The number of PSM and the Mascot score of the SAA2 precursor were ranked fifth and seventh, respectively, among the identified precursors. In contrast, with our quantification method, the highest A/NA enrichment among the amyloid precursors was SAA2 ratio (A/NA=19.6) (**Figure 3c**). Another example is case 6. Here, unlike the previous case, IHC staining was in favor of Lambda chain déposit. Lambda light chain amyloidosis was also suspected on the basis of the patient’s clinical profile (IgG Lambda monoclonal gammopathy) (**Figure 4a**). Amyloidosis-associated proteins (ApoE, ApoA4 and APCS) were identified and were enriched in the deposit (**Figure 4b**). Lambda light chain fragments (IGL) did not have the highest Mascot score or the highest number of PSMs (3rd and 6th among amyloid precursors, respectively). In contrast, the highest enrichment ratio among the precursors was a fragment of the variable domain of IGL (IGLV3-9 ratio A/NA=5.5) (**Figure 4c**). As the highest Mascot score was an IGKC, this erroneous ranking could have led to a different management of the patient. These two examples illustrate the gain in robustness in quantifying protein enrichment in amyloid deposition compared to a simple identification approach.

**Figure 1:**
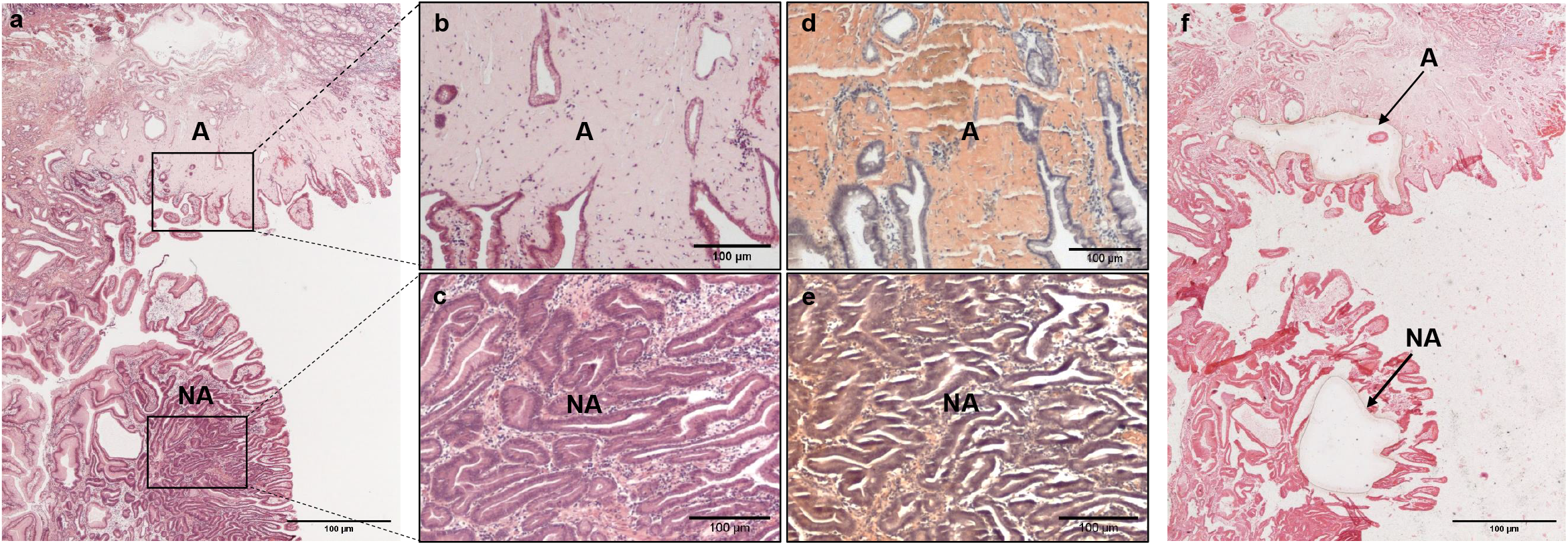
Example of a laser capture of amyloid (A) and non-amyloid (NA) tissue areas in case number 4. Overall hematein eosin saffron (HES) staining from a surgical resection for ampulloma (a). Focus on amyloid deposits seen in the lamina propria (b) and on a non-amyloid area (c) with HES staining, and with Congo red staining (d and e). f-HES section after laser capture. The arrows indicate the microdissected areas. Scale bar = 100 μm.

**Figure 2:**
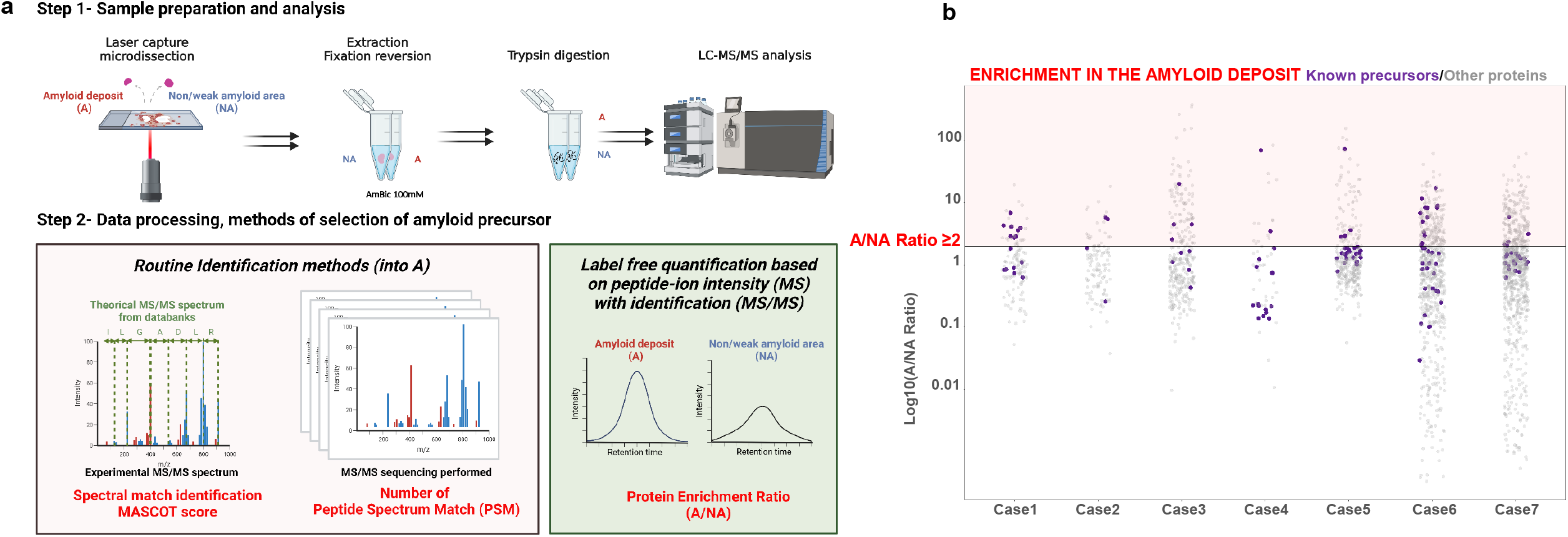
a- Analytical workflow. Step 1 - Microdissection of an amyloid (A) and a non-amyloid (NA) area. Proteins were extracted, digested with trypsin and peptides analyzed by high resolution tandem mass spectrometry (LC-MS/MS). Step 2 - Processing of mass spectrometry (MS) data, the two methods currently practiced in clinical routine are based on the quality of identification from an amyloid area (Mascot score and Peptide Spectrum Match/PSM number). The proposed new method (in green) integrates a quantification of the relative abundances of proteins to quantify an enrichment ratio in the amyloid deposit compared to the non-amyloid area (A/NA). b-Global representation of the identified and quantified proteins for the 7 analyzed cases. Each point represents a protein distributed according to its A/NA ratio. Proteins above the bar have an A/NA ratio greater than or equal to 2 and therefore the protein is enriched in the amyloid deposit. Proteins colored in purple are known precursors of amyloidosis.

**Figure 3:**
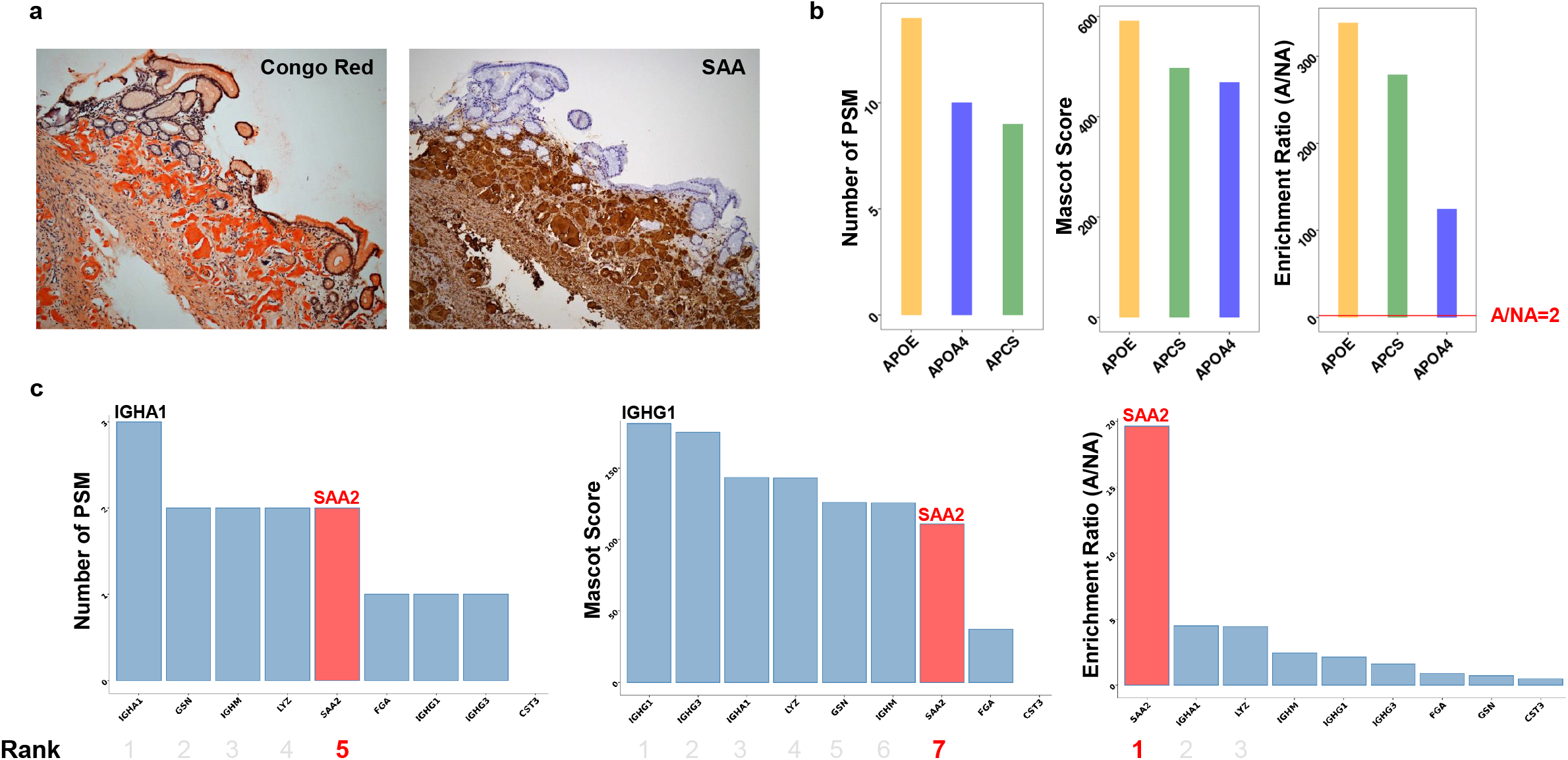
Graphical representation of the results of case 3. **a**- Congo Red staining and Serum amyloid A (SAA) immunostaining that allowed amyloidosis typing in clinical practice. **b-** Identification and quantification of the enrichment of amyloid-associated proteins. At least two of the three must be found enriched to confirm the presence of amyloid deposits in the sample. Enriched proteins have a relative abundance ratio (amyloid area (A) to non-amyloid area (NA) far greater than 2 (red bar). **c-** Ranking results according to the 2 clinical routine identification methods (Mascot score and number of PSM) and ranking of enrichment ratios in the amyloid deposit. The protein associated with the known amyloidosis type is colored in red and its ranking is indicated below. The enrichment ratio is of nearly 20 for serum amyloid A, while other described amyloid precursors only showed a comparatively mild enrichment.

**Figure 4:**
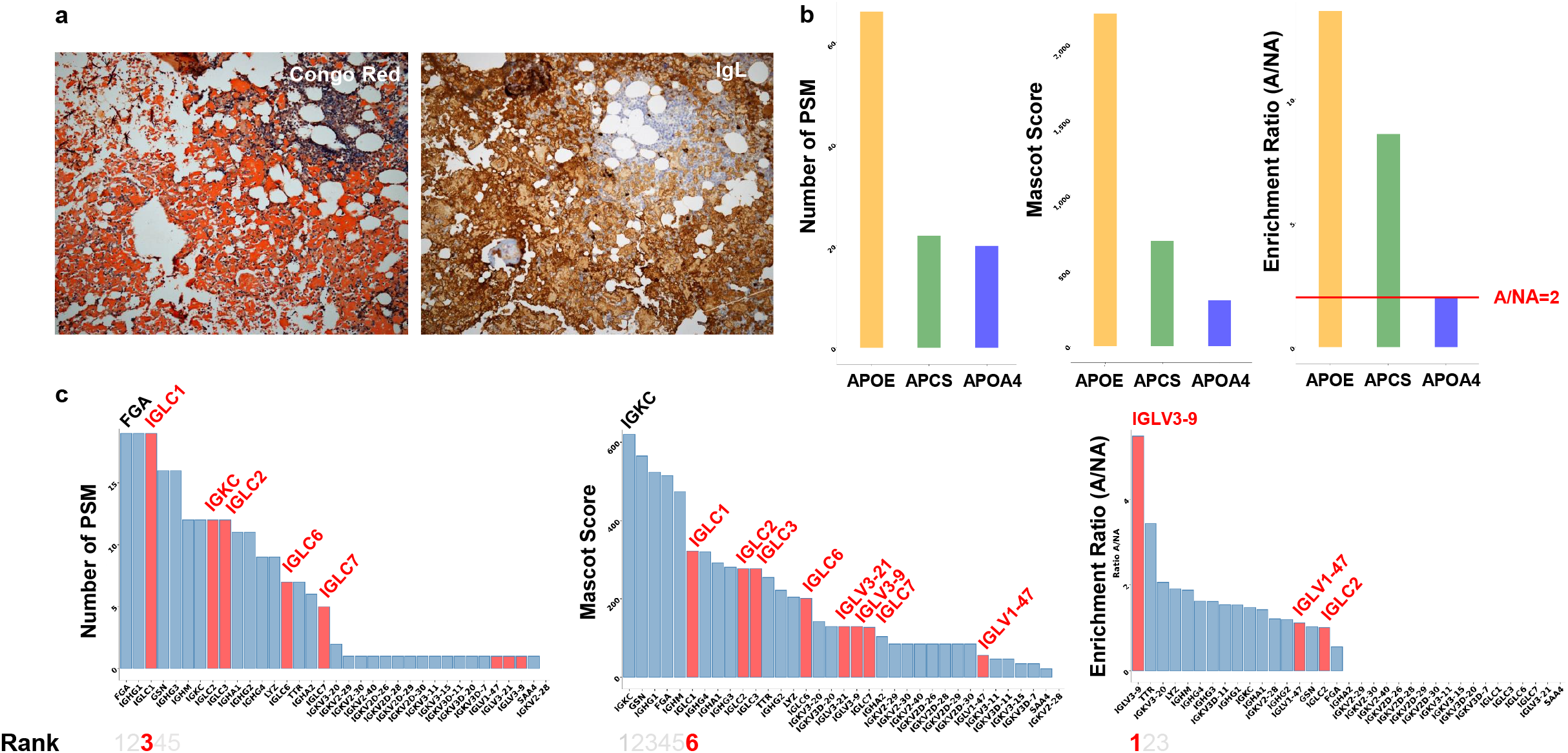
Graphical representation of the results of case 6. a- Congo Red staining, showing large amounts of amyloid deposits in this bone marrow biopsy and Immunoglobulin Lambda (IgL) positive immunostaining b- Identification and quantification of the enrichment of amyloid-associated proteins. At least two of the three must be found enriched to confirm the presence of amyloid deposits in the sample. Enriched proteins have a relative abundance ratio (amyloid area (A) to non-amyloid area (NA) greater than or equal to 2 (red bar). c- Ranking results according to the 2 clinical routine identification methods (Mascot score and number of PSM) and ranking of enrichment ratios in the amyloid deposit among known non-localized precursors. The protein associated with the amyloidosis type are colored in red and the first ranking is indicated below. The enrichment ratio is the highest with a fragment of the variable domain of a lambda light chain, matching the known monoclonal gammopathy of this patient.

Overall, identification and quantification results of the 7 cases analyzed are summarized in **Table 2**. Proteins known to be associated with amyloidosis were systematically identified, with varying levels of enrichment in each case. We found a significant and positive correlation between the APOE Mascot score and the semi-quantitative histological abundance of amyloid deposits (ρ=0.93, p=0.008). No significant correlation was observed with other amyloidosis-associated proteins (number of PSMs or Mascot score), probably due to a lack of statistical power in this series: APCS (ρ=0.20, p=0.66 and ρ=0.45, p=0.31, respectively), APOA4 (ρ=0.52, p=0.29 and ρ=0.52, p=0.29, respectively), APOE (ρ=0.77, p=0.07 for PSM). As for the abundance ratio, no clear correlation was found between the histological amount of amyloid deposits and APOA4 (ρ=-0.46, p=0.36), APCS (ρ=-0.24, p=0.60) or APOE (ρ=0.15, p=0.77). Considering amyloidosis typing, the PSM number-based method retained a correct amyloid precursor in 3 cases out of 7, while results were equivocal in 4. The Mascot score-based method retained a correct amyloid precursor in 2 cases, while results were equivocal in 5. Enrichment quantification retained a correct amyloid precursor in all cases (**Table 2**).

**Table 2:** Identification and quantification proteomic results

| Case No. | Amount of amyloid deposits | Proteins (Peptide spectrum matches, Mascot score, Ratio Amyloid/non-Amyloid) |  |  |  |  |  |  |  |  | Amyloid subtype by IHC | Amyloid subtype by PSM | Amyloid subtype by Mascot | Amyloid subtype (Ratio) |
| --- | --- | --- | --- | --- | --- | --- | --- | --- | --- | --- | --- | --- | --- | --- |
|  |  | Common protein |  |  | Precursors |  |  |  |  |  |  |  |  |  |
|  |  | APCS | APOE | APOA4 | SAA | TTR | B2M | Lambda light chain | Kappa light chain | Other |  |  |  |  |
| 1 | ++++ | 3/3/152/2.8 |  | 3/3/131/12<br>.1 |  |  | 3/3/113/4.<br>9 |  | IGKC<br>2/2/77/3.3 | IGHG1 5/5/234/0.8<br>LYZ 4/5/239/0.6<br>IGHA2 1/1/37/1.6 | Aβ2M | Equivocal | Equivocal | Aβ2M |
| 2 | ++ | 4/4/150/8.1 | 2/2/76/6.3 |  |  |  |  | IGLC2<br>2/2/102/6.6 |  | IGHG2 2/2/96/4.5<br>IGHA1 2/2/104/2.2 | AL Lambda | Equivocal | Equivocal | AL Lambda |
| 3 | +++ | 9/9/498/279 | 12/14/592/339 | 9/10/469/1<br>25 | SAA2<br>2/2/111/<br>19.6 |  |  |  |  | IGHG1 4/4/181/2.2<br>IGHG3 4/4/175/1.6<br>IGHA1 3/3/144/4.5<br>LYZ 2/2/143/4.5 | AA | Equivocal | Equivocal (AH ?) | AA |
| 4 | +++++ | 4/5/242/5.8 | 20/26/1214/18<br>5 | 27/33/141<br>0/115 |  |  |  | IGCL2<br>3/5/186/0.6 | IGKC<br>4/4/344/0.2 | IGHM 10/15/562/61.8<br>FGA 6/6/383/1.9<br>IGHG3 7/10/301/2.6<br>APOA1 7/9/327/31.8 | Non-informative | AH IgM | AH IgM | AH IgM |
| 5 | +++ | 11/45/663/45.5 | 16/30/1138/11<br>8.2 | 20/23/105<br>1/104.2 |  | 9/26/735/94.<br>2 |  | IGLV1-47<br>1/1/77/2.4 | IGKV3-20<br>2/3/174/4.6 | EFEMP1 3/4/259/28.0<br>APOA1 14/17/753/0.9<br>IGHG3 10/16/508/2.5<br>IGHG1 10/16/562/1.3<br>GSN 10/12/706/0.8 | ATTR | ATTR | Equivocal | ATTR |
| 6 | +++++ | 11/23/675/8.6 | 28/68/1816/13.<br>6 | 10/15/515/<br>2.0 |  | 4/6/259/3.4 |  | IGLV3-9<br>1/1/129/5.5 | IGKV3-20<br>2/3/168/2.1 | APOC3 1/1/101/7.7<br>LYZ 4/8/257/1.9 | Non-informative | Equivocal | Equivocal | AL Lambda |
|  |  |  |  |  |  |  |  |  |  | FGA 13/26/813/0.6<br>IGHG1 10/23/610/1.5<br>GSN 12/20/784/1.0 |  |  |  |  |
| 7 | +++++ | 10/12/614/40.5 | 25/41/1624/50.<br>**8** | 30/45/171<br>3/43.7 |  | 8/12/549/3.1 |  | IGLV3-9<br>1/1/152/0.9<br>IGLC2<br>3/5/188/0.44 | IGKV6D-21<br>3/7/240/15.5<br>IGKC<br>9/35/768/6.5 | APOC3 2/2/164/13.4<br>APOA1<br>26/49/1431/11.9 | Non-informative | AL<br>Kappa* | AL Kappa* | AL Kappa |

The amount of amyloid deposits was semi-quantitatively assessed in a + to +++++ scale where +++++ represents a sample with nearly a 100% of amyloid deposits. For each case, results from the common proteins found in amyloid deposits, are firstly displayed, followed by the results for the top precursors of amyloidosis. For each protein, the number of peptide spectrum matches (PSM number of detected MS/MS spectra) is displayed, followed by the Mascot identification score and finally the enrichment in the amyloid area assessed by label free enrichment quantification (abundance ratio between the amyloid microdissected area and the non-amyloid area, highlighted using bold fonts). Interpretations are provided for each approach, as well as the immunohistochemical findings. For each approach, the precursor with the best score was considered as the amyloid subtype (except for APOA1, commonly seen in amyloid deposits). An equivocal result was considered when the second best precursor score was in a 10% range from the first best precursor score (including ties). Abbreviations: Ig, immunoglobulin; IHC, immunohistochemistry; PSM, peptide spectrum matches; XIC, extracted ion chromatogram.

At this stage, we wanted to define an indicator allowing us to compare the different approaches based on mass spectrometry. Whatever the approach used, from all identified and/or quantified proteins, a protein precursor with a top score will emerge (number of PSM, Mascot score or enrichment ratio). Confidence in results interpretation can be summarized, for each approach, as the relative difference between the hit protein compared to the other proteins. We defined a confidence score for each approach calculated by dividing the value associated with the hit protein by the average value of the next 2 proteins. In this way, label free quantification of enrichment in this cohort showed the most reliable results compared to PSM- and Mascot-based approaches (Mann-Whitney U tests, p=0.01 and p=0.007 respectively, **Supplemental Figure 1**). Interestingly, a positive correlation was found between the abundance of histological deposits and the Mascot score (ρ=0.88, p=0.009), whereas this was not the case with enrichment quantification (ρ=-0.26, p=0.61) and doubtful with the PSM (ρ=0.69, p=0.08). This last point suggests that quantification of deposits provides a reliable rationale for identifying the precursor even if deposits are low.

In the end, the contribution of quantification in the selection of the amyloid precursor by mass spectrometry allows to avoid any ambiguity and even if the deposits are small.

## Discussion

Amyloidosis typing is crucial to determine the best therapeutic strategy for each patient. Routine histological techniques like immunohistochemistry on FFPE are frequently challenging or non-informative, due to either background noise, lack of available specific antibodies or lack of remaining material when multiple immunostains are necessary. Hence, laser microdissection of amyloid deposits combined with tandem mass spectrometry has emerged as the new gold standard for amyloidosis typing when routine techniques have failed. The two main proteomics workflows described in the literature for protein precursor selection are based on the stratification of identified proteins according to the Mascot identification score or the number of PSM. However, data interpretation is sometimes a challenge, particularly when multiple protein precursors are identified, possibly be due to a lack of quantification. While being considered the new gold standard of amyloidosis typing, only few centers in the world have currently integrated proteomic analysis based on mass spectrometry in their clinical routine. Most of those who have mastered this technique have acknowledged difficult cases where multiple precursors are detected with the same identification quality [7,14,24,25]. In these studies, amyloid precursor selection was based on either the Mascot identification score [7] or the number of PSM [14,24,26,27], primarily from a single microdissection of amyloid area. The amyloid precursor with the best Mascot score or the highest number of PSM is selected as a ground rule. These methods have the advantage of an ultra-simplified interpretation of results without extensive bioinformatics processing. However, these scores are limited to a protein identification quality and have no real quantification value.

Here we introduce label free enrichment quantification for amyloid precursor selection and highlight its usefulness in difficult cases of amyloid typing. Indeed, one of the simplest solutions to discriminate the precursor among several identified proteins remains the quantification of the enrichment between the deposit and an additional non-amyloid area. In addition, Mascot score and PSM number-based approaches of amyloidosis typing may be more difficult in cases where deposits are less abundant, as illustrated in the following works [11,28], whereas enrichment quantification remains robust even with lower ratios.

We noticed that the number of PSM was overall lower in our analyses compared to those of the literature. This is probably due to a different time value of dynamic exclusion in our methods, but we did not have access to this information in the literature. In addition, in our study we worked from microdissected amyloid and non-amyloid areas of 1 mm^2^, comparing for each patient always the same amount of tissue between the deposit and the non-amyloid area. This surface range is higher compared to some scarce amyloid deposits encountered in routine practice and the common workflow described by the Mayo Clinic and the UK National Amyloidosis Centre. Our approach considers that it is better to interpret a deeper proteome in a quantified way than to limit oneself to the identification, ultimately enabling to increase the chances of a correct precursor selection. By taking a slightly larger amount of tissue, it doesn’t matter if other potential precursors are identified because the enrichment ratios will be different and will allow the right selection.

The results of the abundance ratio show the best robustness from our series as compared to other published workflows. The enrichment quantification highlighted the expected protein precursor as the main quantified protein enriched in amyloid deposits among the candidate precursors in all 7 cases, independent of the amount of histological deposits. In contrast, the Mascot and PSM-based approaches had difficulties in about half of the cases, with a significant drop in confidence when amyloid deposits were scarce. Indeed, by defining a confidence score for amyloid precursor selection, we showed that enrichment quantification provided a more reliable choice than Mascot and PSM-based approaches in this series. This was particularly visible in our cases 3 to 5, where enrichment quantification allowed a greater confidence in the identification of the amyloid precursor.

Of course, in this study, we only tested our approach on a small cohort of 7 cases. Still our goal to report a proof-of-concept of the usefulness of label free enrichment quantification is achieved. Beyond the percentage of successful amyloidosis typing, the prospect of analyzing large collections with label free quantification of amyloid precursor enrichment could potentially revise the reliability of methods currently used in clinical routine. Here we quantify a protein enrichment, which requires the presence of non-amyloid tissue as control. If there is none, a less amyloid area can serve as a control and will be sufficient to quantify an enrichment in the deposits.

It is also interesting to notice that several precursor proteins can be enriched in an amyloid deposit and that other proteins that are not known to be related to amyloidosis can also be enriched. Proteomic profiling of amyloid deposits in a large collection of patients could undoubtedly allow a better characterization of the pathology and the identification of new precursors, the number of which continues to increase [3,4].

To conclude, this study brings a proof-of-concept of the usefulness of label free enrichment quantification as a valuable improvement for amyloidosis typing by mass spectrometry-based proteomics. We highlighted that label free enrichment quantification can significantly ease amyloid precursor selection, at the reasonable price of analyzing a supplementary non-amyloid area and requiring further bioinformatics expertise. Quantification of enrichment is a readily applicable method. It can be used as a first-line approach to amyloidosis typing, which will avoid equivocal diagnosis and improve patient management.

## Data Availability

The mass spectrometry proteomics data have been deposited to the ProteomeXchange Consortium via the PRIDE partner repository with the dataset identifier PXD039814.

## Abbreviations

A: Amyloid
FDR: False Discovery Rate
FFPE: formalin-fixed and paraffin-embedded
Ig: immunoglobulin
IHC: immunohistochemistry
LC-MS/MS: liquid chromatography and tandem mass spectrometry
NA: Non-amyloid
PSM: peptide spectrum matches
XIC: extracted ion chromatogram

## Figure Legends

**Supplemental Figure 1:** The confidence score in interpreting the results was summarized as the relative difference between the three protein precursors with the highest score/ratio for each approach, by dividing the score/ratio of the first precursor divided by the average of the next 2 scores/ratios. A higher confidence score means greater confidence in selecting the protein precursor from the observed data. Mann Whitney U tests were performed to compare the three amyloid precursor ranking methods: Mascot score-based method, number of PSM (peptide spectrum matches)-based method and enrichment quantification (abundance ratio).

