## Supplementary figures and images for "Quantification of amyloid protein enrichment by mass spectrometry improves amyloidosis typing"

### Supplementary Figure 1

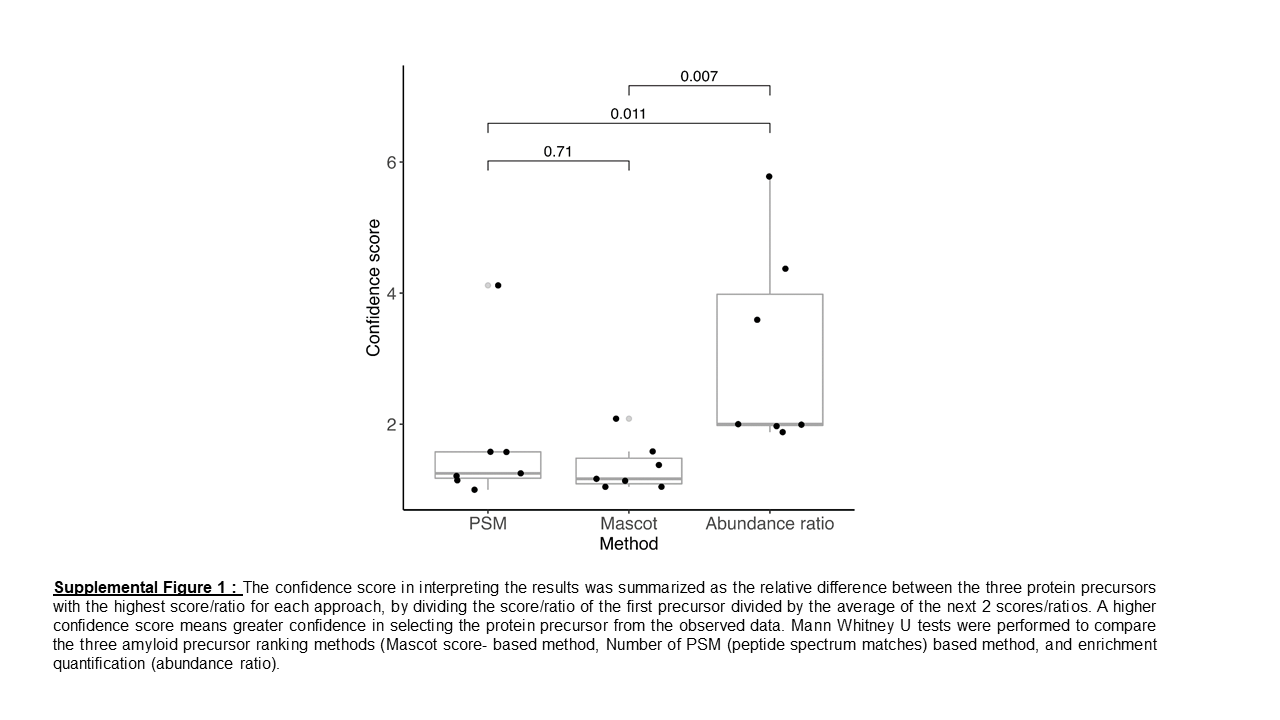
